# Systematic Review and Meta-Analysis on the Effect of Self-Assembling Peptide P_11_-4 on Initial Caries Lesions

**DOI:** 10.1101/2022.06.13.22276212

**Authors:** Jeremy Horst Keeper, Laura J Skaret, Madhuli Thakkar-Samtani, Lisa J. Heaton, Courtney Sutherland, Kathryn Vela, Bennett T. Amaechi, Anahita Jablonski-Momeni, Douglas A Young, Jeanette MacLean, Robert J Weyant, Andrea Ferreira Zandona, Domenick Zero, Woosung Sohn, Nigel Pitts, Julie Frantsve-Hawley

**Author notes:** **Corresponding author.** Address: 465 Medford Street, Boston, MA 02129-1454. **Funding** There is no funding to report related to this work. **Contributions** CareQuest Innovation Partners in collaboration with CareQuest Institute for Oral Health led the development and authorship of the systematic review and meta-analysis in collaboration with an expert panel. The expert panel informed and reviewed all aspects of the study protocol prior to registration. All co-authors voted on each of 23 aspects of the study plan. We discussed items with less than 80% agreement until this level of consensus was reached. We shared and reviewed all included studies, extracted data, and bias interpretations regularly with all co-authors. All co-authors contributed to and reviewed the final manuscript.

## Abstract

**Background:** Dental caries remains a global problem that causes disproportionate suffering in underserved populations. Simple interventions are needed to improve patient experience, clinical and cost-effectiveness, and access to care. The self-assembling peptide P_11_-4 is a recently developed, non-invasive treatment that regenerates enamel in initial caries lesions.

**Studies reviewed:** We conducted a systematic review and meta-analysis on the effectiveness of the P_11_-4 products Curodont™ Repair (CR) and Curodont™ Repair Fluoride Plus (CRFP) on initial caries lesions. Primary outcomes were lesion progression after 24 months, caries arrest, and cavitation. Secondary outcomes were changes in merged International Caries Detection and Assessment System (ICDAS) score, Quantitative Light Fluorescence, esthetic appearance, and lesion size.

**Results:** Six clinical trials comparing CR to controls met the inclusion criteria. Results of this review represent two primary and two secondary outcomes. When compared to parallel groups, CR improved caries arrest (RR: 1.82; 95% CI: 1.32 to 2.50; 45% attributable risk; 95% CI: 24 to 60%) and decreased lesion size by 32% (Hedge’s g: -0.59; CI: -1.03 to -0.15). We observed positive trends for avoiding cavitation (RR: 0.32; CI: 0.10 to 1.06) and lowering merged ICDAS score (RR: 3.68; CI: 0.42 to 32.3). No studies used CRFP or reported adverse esthetic changes.

**Practical implications:** CR has a treatment effect on caries arrest and decreased lesion size. Two trials contributing to the caries arrest result had non-masked assessors, and all trials had elevated risks of bias. We recommend conducting longer trials. CR is a promising treatment for initial caries lesions.

## Introduction

Globally, dental caries (tooth decay) remains a widespread public health problem, with untreated cavitated caries lesions being the most common condition in the *Global Burden of Disease Studies*.^1–3^ This disease causes disproportionate suffering in underserved populations.^4–6^ The reliance of traditional oral healthcare on invasive, treatment-sensitive treatments requiring extensive training results in a system where fear and expense are barriers to adequate oral healthcare access for large portions of the population.^3,7,8^ Effective and simple non-invasive interventions are needed to improve the efficacy and efficiency of care and to empower upstream policy interventions.^3,9,10^

Unlike most tissues in the body, tooth enamel cannot regenerate once a hole (actual cavity) has formed. Dental caries lesions form when the minerals of the tooth are dissolved out (demineralization) due to dental biofilm bacteria (plaque) fermenting dietary sugars (processed carbohydrates) into acids that diffuse into the tooth. Demineralization results in weakened and porous tooth structure. The first stage is often misleadingly described in the United States (US) as a “cavity,” though it is not cavitated (there is no hole); international and American Dental Association (ADA) terminologies employ the term *initial caries lesion*.^11–14^ A tooth can have initial lesions without cavitation because of the phasic nature of demineralization and remineralization,^15^ combined with the hierarchical microstructure of enamel: prismatic enamel rod hydroxyapatite is much more resilient to acid demineralization than interprismatic enamel. As a result, the area is affected but the overall structure remains. Saliva protects and heals initial lesions through physical cleansing, acid pH buffering, and remineralization via calcium, phosphate, and other molecules. The caries lesion progresses when there is not enough protection to offset sugary diets and caries-mediating bacteria.

As the demineralization of the enamel and outer dentin progresses, the outer surface collapses, resulting in a cavitation that allows bacteria into the dentin. Without intervention this usually leads to pain and infection. Traditional treatment involves expensive and technique-sensitive dental operative procedures to restore the damaged tooth structure, for example a dental filling or crown. With continued consumption of sugars and imperfect dental materials, the margin where the filling material meets the tooth may break down through the same caries process, and the area is retreated with a larger filling or crown. This process is cyclic and progressive, leading to more invasive dental or surgical treatment, expense, and suffering.^3,16,17^

Interventions are available to halt (arrest) the caries process. Improved nutrition is paramount.^9^ Nearly all treatments recommended by the ADA to arrest initial caries lesions work through the effects of fluoride^18^ and/or by physically sealing the area (dental sealants and resin infiltration).^19^ The ADA also recommends arresting cavitated lesions by silver diamine fluoride (SDF).^18^ Because cavitated lesions are more difficult to arrest due to being infected and less cleansable, it is expected that initial lesions will be arrested with SDF. There is some evidence to support this: a case series^20^ and one randomized split-mouth study both document lower rates of caries progression than expected.^21^ While these approaches are effective in arresting initial caries lesions, each has limitations. Short-term patient experience can be a barrier, as SDF and traditional fluorides can have aversive taste and SDF stains porous tooth structure black.^22–25^ Dental sealants are an option, but usually only for pits and fissures, and loss of resin-based sealants is associated with the risk of developing caries.^26^ Additional non-invasive therapies without these side effects are sought to complement the existing options.

Relatively new to this field, P_11_-4 is a self-assembling peptide in a brush-on liquid, applied after cleaning and chemical preparation, that works by guiding and catalyzing the regeneration of lost enamel in an initial, non-cavitated lesion. The P_11_-4 peptide, also called Curodont™ Repair (CR), has the amino acid sequence QQRFEWEFEQQ. It is kept separate as a lyophilized powder and rehydrated prior to application. The mechanism is as follows: peptides absorb into an initial lesion, where they self-assemble into long structures inside the lesion, like the rungs of a ladder. This scaffold attracts and integrates calcium, phosphate, and hydroxyl ions into hydroxyapatite. This guided remineralization process for regenerating damaged enamel shows strong results in two weeks in laboratory settings,^27^ and studies show this process promotes bone formation by the same mechanism.^28^ Research provides evidence that CR is clinically safe.^29^ CRFP is registered with the US Food and Drug Administration (NDC 72247-101) as an anticaries drug under the fluoride monograph (21CFR355). There is no stain or taste.

Since its introduction, there have been numerous clinical studies of CR. However, the efficacy of CR for the treatment of initial caries lesions has not been established through systematic reviews or meta-analysis. Therefore, we conducted this systematic review to assess the efficacy across multiple clinical outcomes between CR and control procedures for treating initial caries lesions.

## Methods

### Protocol registration

The protocol for this systematic review was registered *a priori* with PROSPERO (304794).

### Inclusion criteria

This systematic review includes randomized controlled clinical trials and follows the methodology from the Cochrane Review Manual^30^ with minor modifications described below. We evaluated full-text reports identified from screening based on the inclusion and exclusion criteria developed by the ADA Center for Evidence-Based Dentistry in the systematic review^31^ for the ADA clinical practice guidelines on non-restorative treatment for caries lesions.^18^ For inclusion in this review, studies met the following criteria:

*Participants*: individuals of any age with initial (non-cavitated) caries lesions in at least one permanent tooth;

*Intervention*: application of topical CR or Curodont™ Repair Fluoride Plus (CRFP);

*Comparisons*: placebo, fluoride varnish, or no intervention;

### Outcomes

The primary outcomes were caries arrest assessed via visual-tactile methods; cavitation (including restoration); and caries progression after at least 24 months. The secondary outcomes were decrease in International Caries Detection and Assessment System^12,13^ score, with scores 1-2, 3-4, 5-6 merged as in the ADA caries classification system^11^ (merged ICDAS); Quantitative Light Fluorescence^32^ (excluding other quantitative methods using light, fluorescence or thermography, e.g., DIAGNOdent™ or the Canary System^®^); lesion size by radiography or digital photography; and esthetic appearance including discoloration or stain (including report as an adverse outcome). We accepted assessment of primary and secondary outcomes at any time point except for caries lesion progression, for which we required at least 24 months.

### Exclusion criteria (adapted from ADA guideline)

not reporting outcomes on lesions existing at baseline (incidence); not a peer-reviewed paper; randomization method not described; not reporting caries activity by numbers of lesions; not reporting baseline caries status; not reporting product description by brand or concentration; articles not published in English.

### Literature search

A medical librarian (KV) developed a search strategy for English language papers. We performed the search strategy in PubMed with the following query: (self-assembling peptide OR Curodont OR “P(11)-4” OR “P11-4” OR CH3CO-QQRFEWEFEQQ-CONH2 OR 72247-101-12 OR “P11-4 peptide”[nm]) AND ((Dental Caries[MeSH Terms]) OR (dental decay OR carious lesion* OR dental white spot* OR white spot lesion* OR WSL OR cavit* OR initial cari* lesion* OR non-cavitated caries OR incipient OR early carious lesion* OR early caries lesion* OR enamel lesion* OR enamel caries lesion* OR incipient carious lesion* OR caries lesion* OR cari*). We performed a corollary search in Embase. We ran searches for all papers published until November 19, 2021.

The team assessed citations within all identified clinical trial papers and reviewed additional studies. We also contacted the manufacturer of Curodont Repair (vVARDIS, Switzerland; previously Credentis, Switzerland) to request knowledge of any additional trials. Two contributors (LS, JHK) independently screened titles and abstracts of studies identified from the search in duplicate based on the inclusion/exclusion criteria. These contributors also conducted a full text review independently and in duplicate. For both the title/abstract review and the full text review, contributors resolved disagreements by discussion and development of consensus.

### Data extraction

Two contributors (LS, JHK) independently extracted data in duplicate from included studies using specially developed data extraction forms. They collected data regarding primary and secondary outcomes; baseline caries activity status; diagnostic criteria and methodology; randomization and allocation process; adherence to allocation; missing data; measurement methodology; masking of assessors (*blinding* [sic]); reporting for risks of bias; intervention details for all groups; background exposure (e.g., fluoride); and adverse outcomes.

For three trials, we contacted study authors who provided separated outcome data for lesions reported as active by visual-tactile criteria at baseline.^33–35^ For another, we contacted authors who provided raw lesion size data^36^ to make it combinable with the other study reporting this outcome.^35^ We extracted and compared data. The team resolved disagreements by discussion and consensus.

### Risk of bias

Following calibration, two contributors (LS, JHK) independently and in duplicate assessed the risk of bias of each included study against key criteria using the Cochrane Risk of Bias tool 2.0:^37^ randomization, effect of assignment, missing outcome data, measurement, and selective reporting. Authors resolved disagreements by consensus and consulted a third author (JFH) as necessary.

### Data synthesis

We conducted a meta-analysis of studies with combinable results, using Stata version 17 (StataCorp, Texas). We calculated Rrisk ratios (RRs) and confidence intervals (CI) from raw numbers for dichotomous outcomes. We also calculated pooled RR estimates using a random effects model that uses inverse variance methods to assign more weight to larger rather than smaller studies and accounts for both within- and between-study variability.^38^ For continuous outcomes, we calculated summary effect size using the Hedges’ g statistic for standardized mean differences. We estimated heterogeneity using the I^2^ statistic (>50% was considered substantial) and the Cochrane Q test.

### Subgroup and sensitivity analyses

study design (split mouth versus parallel arm); effect of study duration on outcome; immediate treatment with fluoride in experimental and control groups or not; and baseline lesion size by depth, merged ICDAS, physical size by photograph or radiograph, or QLF scores.

## Results

### Search results

We identified one hundred and ninety-three papers from the PubMed (123) and Embase (70) searches, and identified three papers through other sources. Of these, 55 were duplicated studies, resulting in 141 publications. We screened eighteen papers for full text review. We included six studies in the systematic review (Figure 1). Two of the included studies randomized at the patient level,^33,39^ three randomized by side or quadrant (split-mouth),^34–36^ and one randomized various numbers of teeth by pairs within each patient (Table 1).^40^ One study^40^ was not included in the meta-analysis because it did not report any outcome with control and intervention groups.

**Figure 1.**
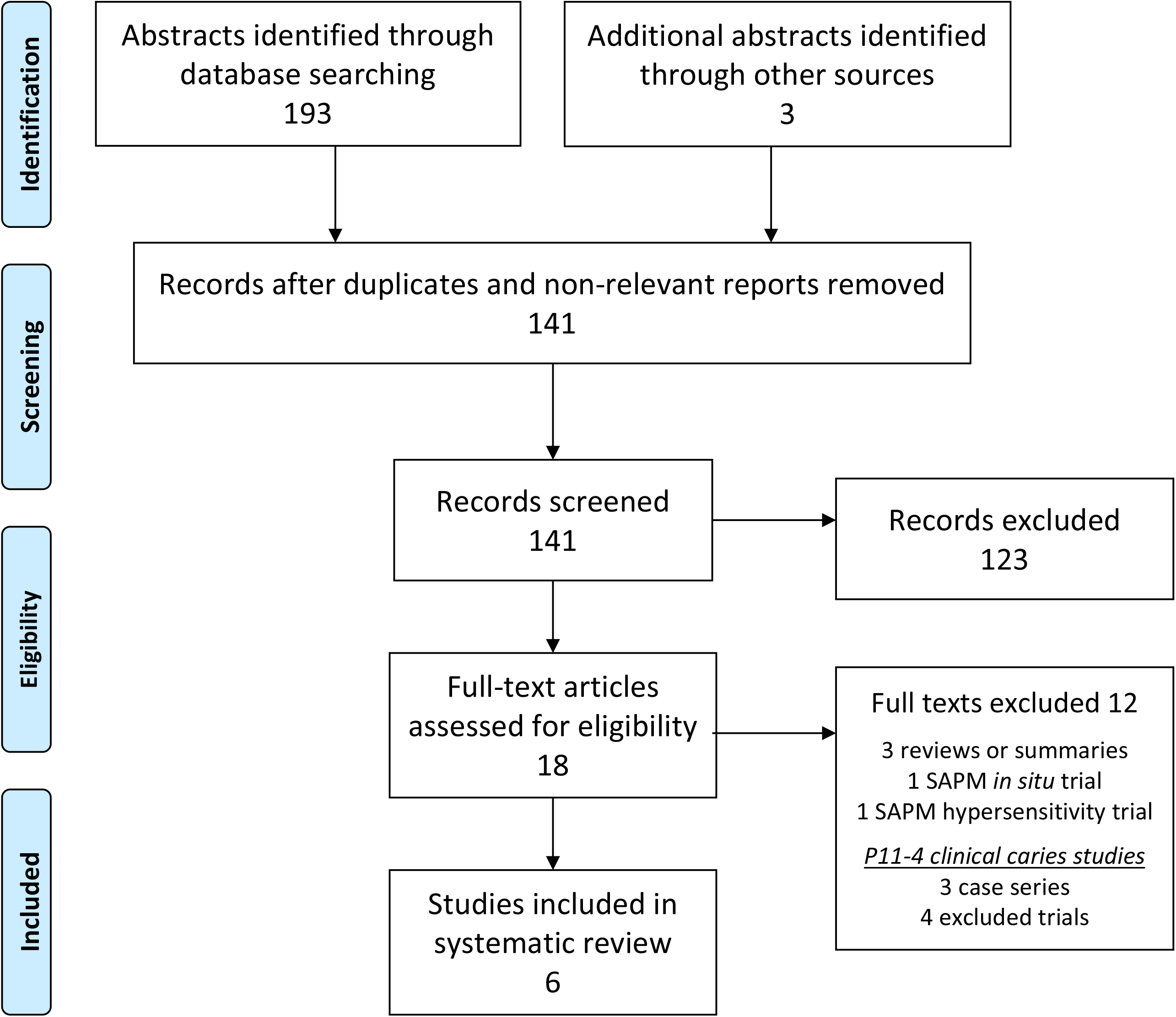
Flow diagram showing the process of identifying, screening, assessing for eligibility, excluding, and including. Abbreviation: SAPM, self-assembled P_11_-4 matrix.

**Table 1.**
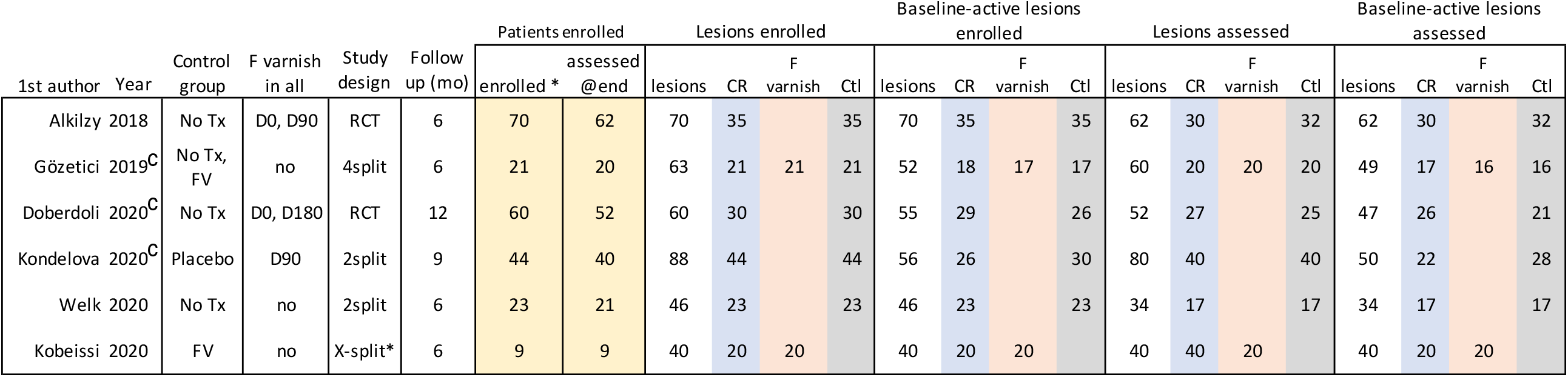
Characteristics, enrollment, and retention of CR clinical trial reports included in this systematic review. Abbreviations: Tx, treatment; FV, fluoride varnish; mo, months; CR, Curodont Repair; Ctl, control; c, contacted study authors and received separate outcomes data for baseline active lesions. *Enrollment excludes treatments not considered in this review; X-split refers to randomized of one or more lesion pairs in each patient.

Five studies were combined in meta-analyses.^33–36,39^ The follow-up length of these 5 studies ranged from 6 to 12 months (mean ± standard deviation: 8±3mo). Studies included participants enrolled into groups of 9 to 70 participants (38±24, sum (Σ): 227). Studies involved between 40-70 active caries lesions (53±10, Σ:319), of which 18-35 were treated with CR (25±6, Σ:151) and compared to a parallel group. In total, we assessed endpoints for 132 lesions active at baseline treated with CR (87% retention) and compared to a parallel group. All six included trials reported the number of lesions that progressed to cavitation (including restoration; Table 2).

**Table 2.**
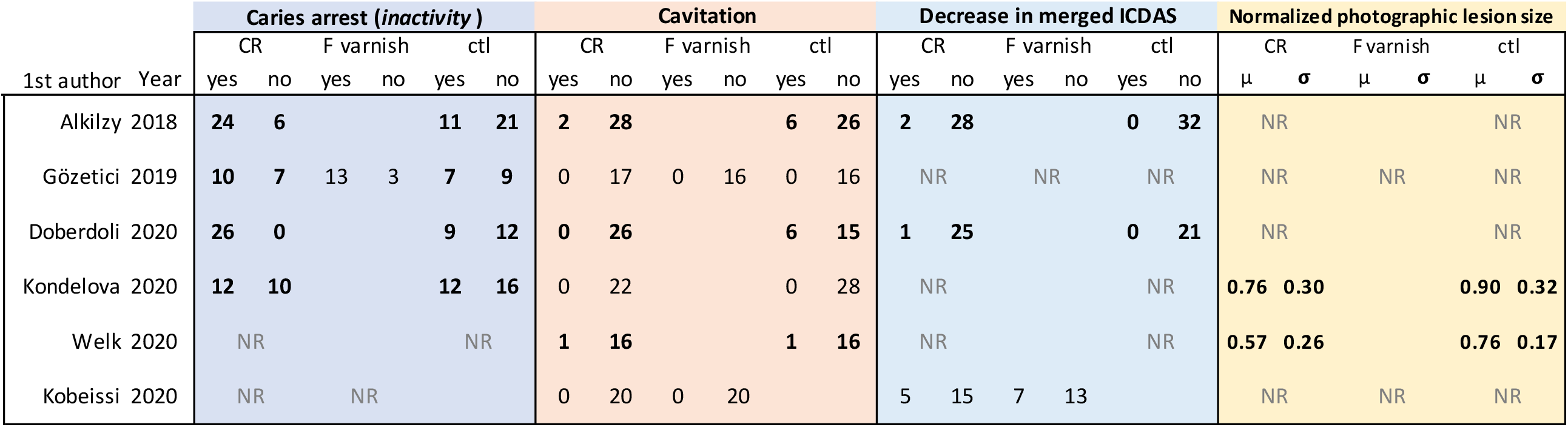
Outcome data for treatment of baseline active initial caries lesions (assessed with visual-tactile systems) with CR, fluoride varnish, and no CR controls for primary and secondary outcomes considered in this review. These values represent a subset of published data for the Gözetici, Doberdoli, and Kondelova studies, which included mixed baseline active and arrested lesions. Abbreviations as in Table 1, and: NR, not reported; µ, mean; ⍰, standard deviation.

### Bias

We assessed overall risk of bias as moderate to high for all studies (Figure 2; Supplementary Table 1). The risk of bias increased due to: 1) lack of masking clinical and statistical personnel, 2) lack of placebo experiences and/or masking patients, 3) lack of prospective public trial registration, 4) missing data, and 5) differences in baseline caries levels between groups.

**Figure 2.**
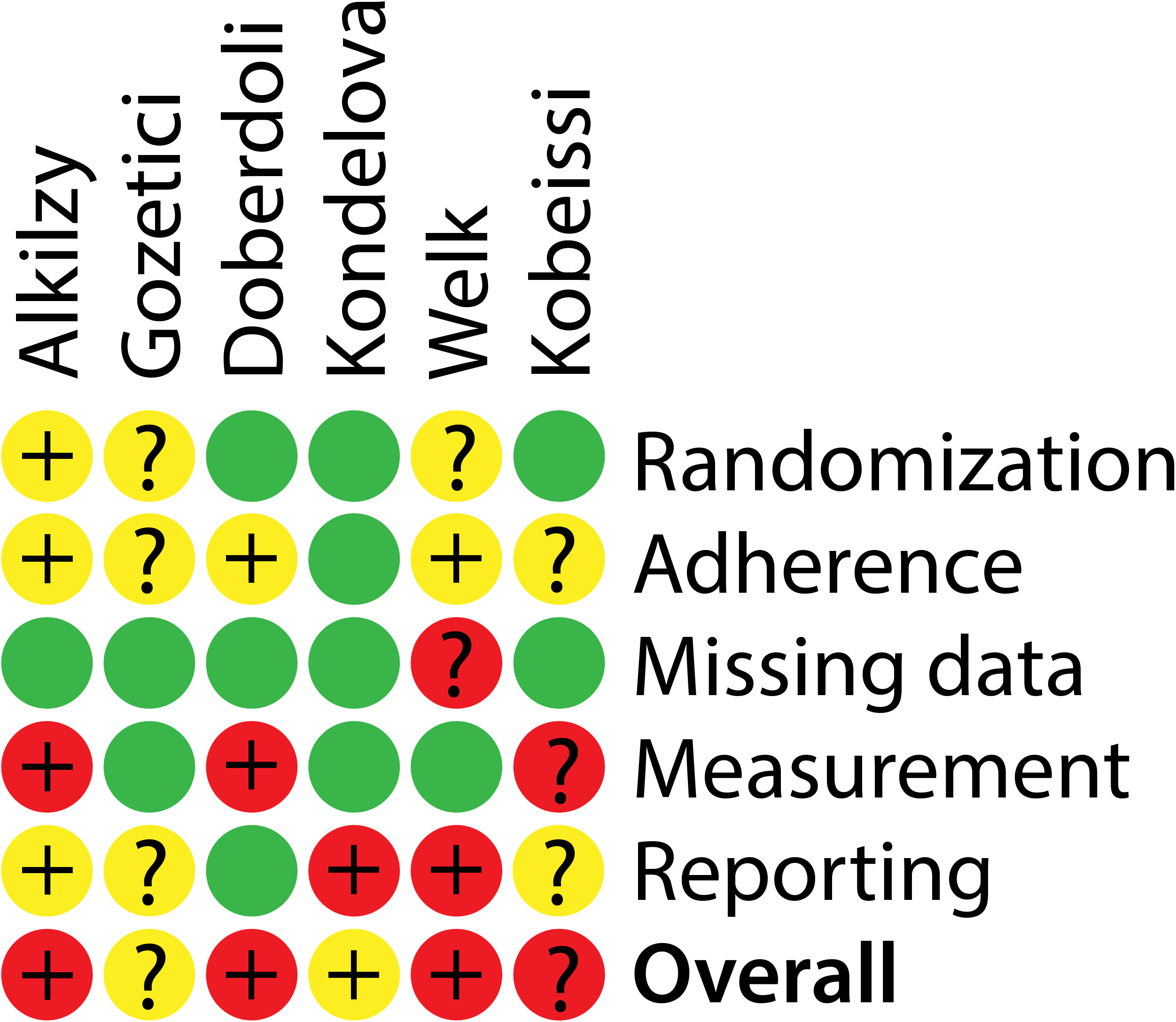
Summary of bias risk across included clinical trials. Details in Supplementary Table 1.

### Primary outcome 1. Effect of CR on caries arrest

Four trials reported caries arrest: three^33,35,39^ used the Nyvad criteria,^41^ while one^34^ used LAA-ICDAS.^42^ Meta-analysis of the four trials reporting on caries arrest found a risk ratio of 1.82 (95% CI: 1.32-2.50; Figure 3). This finding translates by 1-1/RR to an attributable risk^43^ of 45% (95% CI: 24-60%); i.e. the proportion of all teeth treated with CR estimated to arrest due to CR.

**Figure 3.**
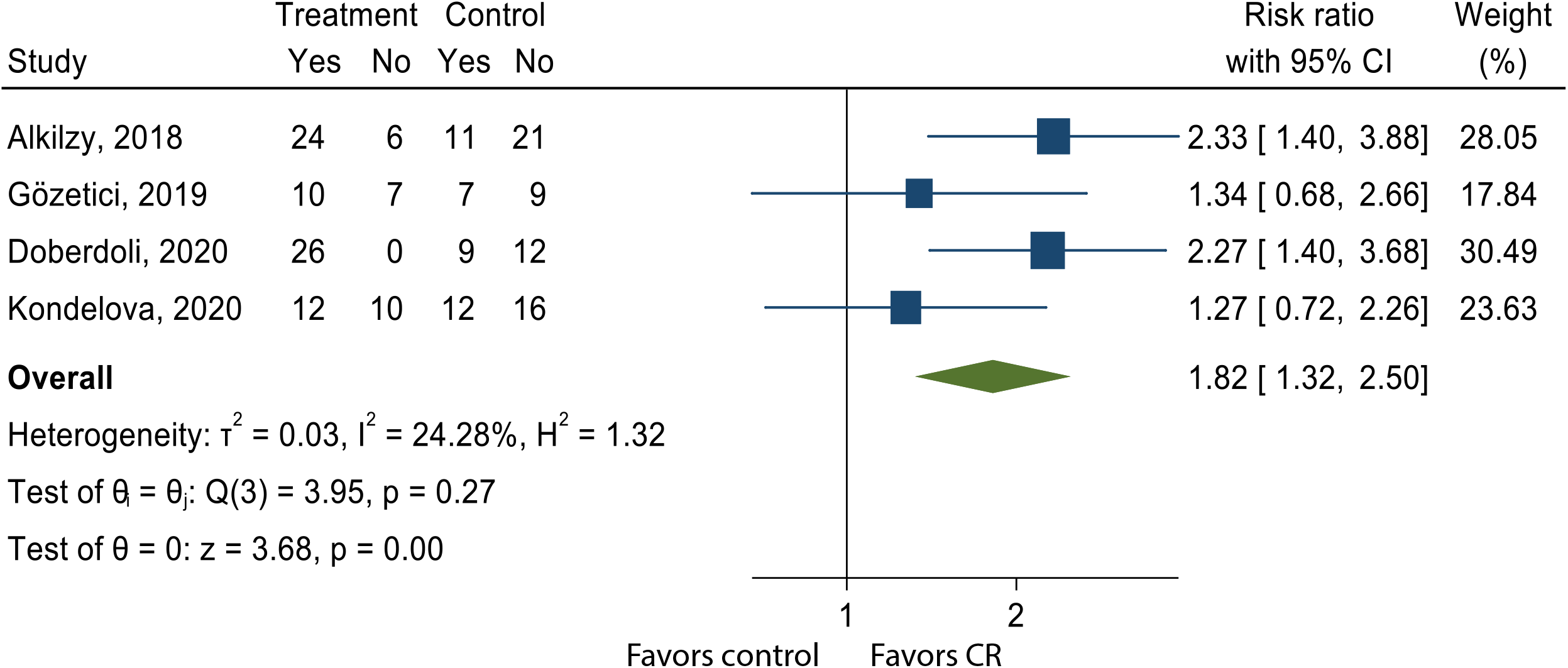
Meta-analysis of caries arrest in randomized-controlled trials of CR shows an effect, estimating that 45% (95% CI: 24-60%) of initial caries lesions that would not arrest naturally, arrest with CR treatment in 6-12 months. The two studies driving this result had non-masked assessment.

Two trials showing the largest effect had non-masked assessment of outcomes,^33,39^ which poses a risk to bias. These two studies are also the only patient-randomized (non-split mouth) studies and the only to combine fluoride varnish with CR during the intervention (Table 1). They are also the only two trials to assess treatment of occlusal surfaces, whereas all four other included trials studied smooth surfaces (non-approximal). Testing for subgroup differences for this division of studies resulted in a p-value of 0.05. (Figure S1).

Testing for subgroup differences in study duration on caries arrest resulted in a p-value of 0.87 (Figure S2).

### Primary outcome 2. Effect of CR on cavitation

All studies reported on cavitation, but only three observed any occurrence; these studies were 6, 6, and 12 months in duration.^33,36,39^ Meta-analysis found a trend for reducing the risk ratio of cavitation (RR=0.32, 95% CI: 0.10-1.06; Figure 4). Lack of cavitation in the other three studies likely include false negative outcomes that would be captured accurately in longer studies.^34,35,40^ Longer studies would be better positioned to assess whether CR prevents cavitation.

**Figure 4.**
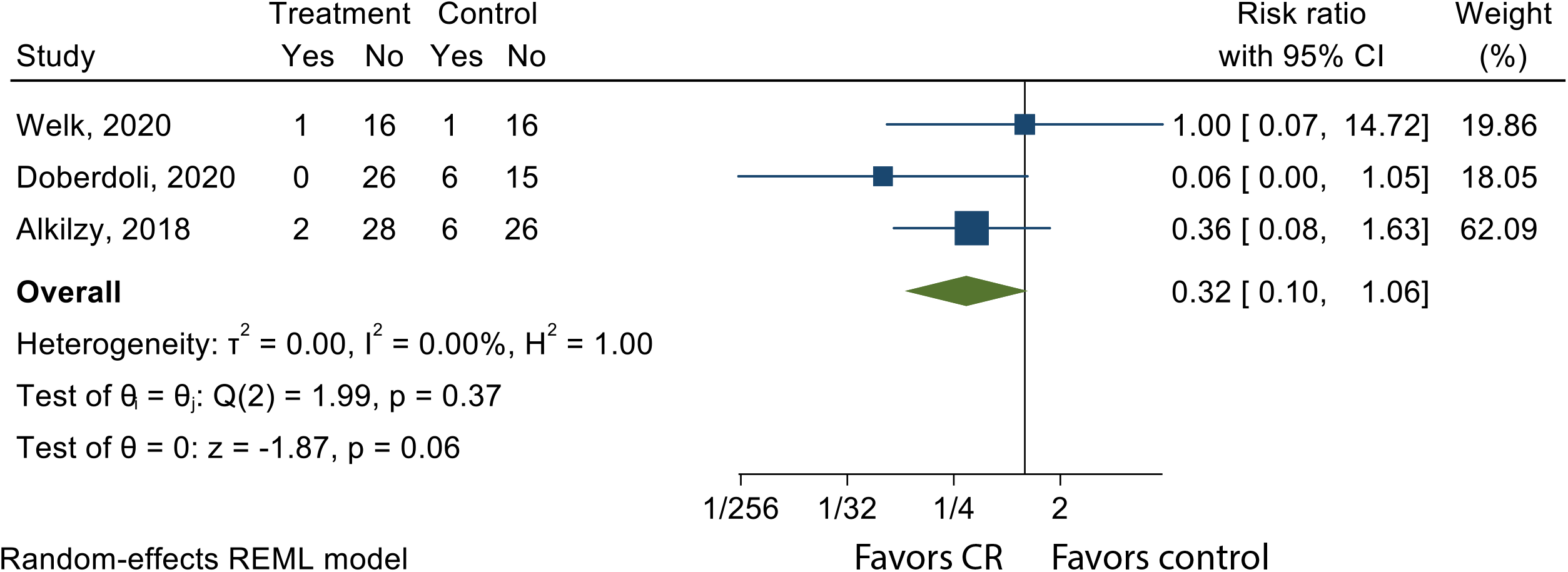
Meta-analysis of preventing cavitation in randomized-controlled trials of CR.

### Secondary outcome 1. Effect of CR on decrease in merged ICDAS

Three trials reported change in ICDAS score^33,39,40^ and were assessed by the merged criteria, with a change from 3 to 0-2 or 1-2 to 0 qualifying as a decrease. We found the risk ratio for decreasing merged ICDAS was 3.68 (95% CI: 0.42-32.26; Figure 5). The large confidence interval occurs due to zero events in the control group. Accordingly, we calculated the risk difference (0.05; 95% CI: -0.01-0.11) as an exploratory analysis (Figure S3).

**Figure 5.**
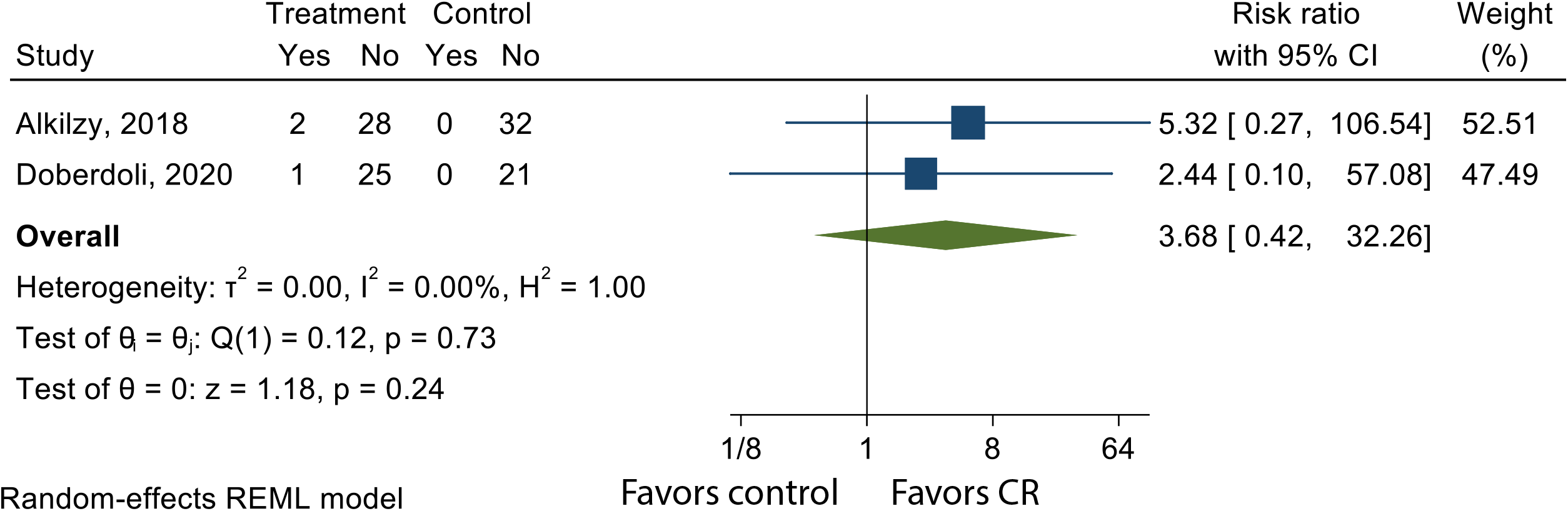
Meta-analyses of decrease in merged-ICDAS score in randomized-controlled trials of CR.

The included study^40^ that does not have groups matching other studies (not represented in meta-analyses) had a risk ratio of 1.15 for CR compared to fluoride varnish for a decrease in merged ICDAS (95% CI: 0.77-1.74).

### Secondary outcome 2. Effect of CR on lesion size

Two trials reported change in lesion size measured by automated software from digital photography.^35,36^ Meta-analysis revealed a standardized mean difference in favor of CR treatment, reducing caries lesion surface size by 32% ± 28% (mean ± standard deviation) with an effect size of -0.59 (Hedge’s g; 95% CI: - 1.03 to -0.15; Figure 6).

**Figure 6.**
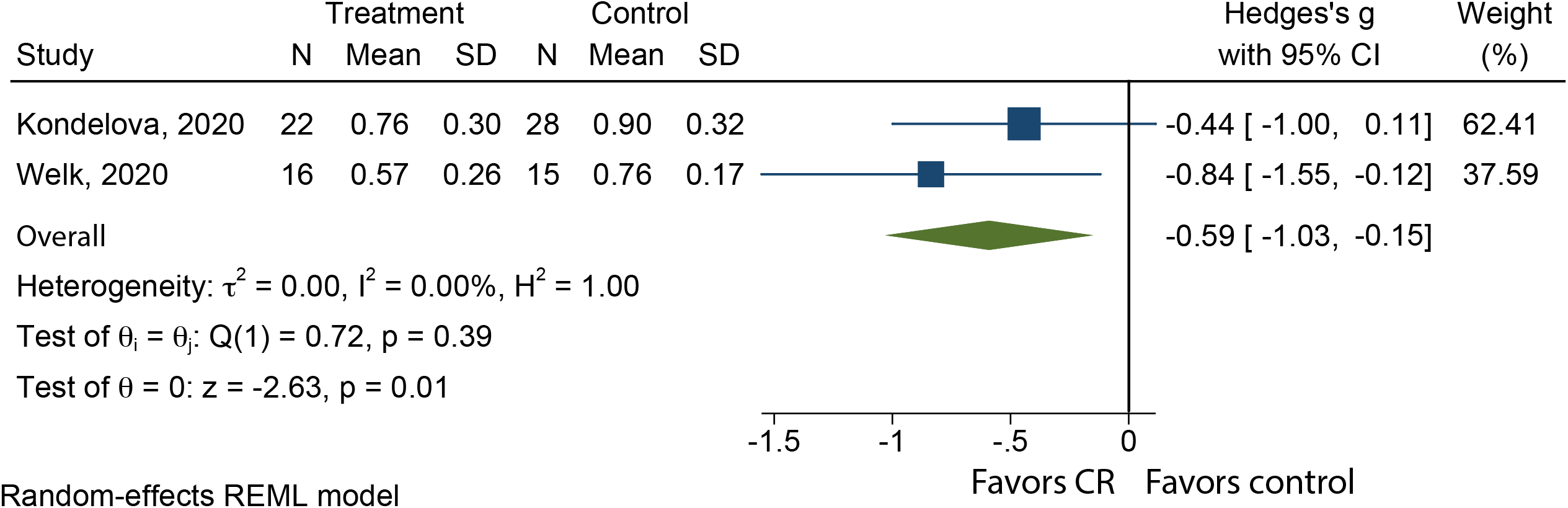
Meta-analyses of decrease in caries lesion surface size in randomized-controlled trials of CR shows an effect.

### Other outcomes

No trials were long enough to measure caries progression. No trials used QLF as an outcome measure. No included trials reported on esthetic appearance, though one excluded trial reported on color change assessed through a spectrophotometer.^44^

### Adverse outcomes

Four included trials reported no adverse outcomes,^33,35,39,40^ two of which explicitly measured them.^35,39^ One excluded case series involving 15 healthy adults reported one patient with dentin hypersensitivity and one with new sensitivity with a chlorhexidine mouth rinse.^29^ No studies reported adverse esthetic changes.

## Discussion

This systematic review and meta-analysis found a potential therapeutic effect for CR on the primary outcome of arresting initial caries lesions in four trials ranging from 6-12 months in duration. Overall, 80% of all caries lesions across the trials were arrested. For 56% of these arrested caries lesions the arrest could be attributed to the effect of CR (44% due to other factors). In other words, considering all teeth, 45% of caries lesions were arrested due to CR, as the risk ratio (RR) of 1.82 translates into an attributable risk fraction of 45% (1-1/RR). Longer studies could show stronger results for prevention of cavitation and regression in merged ICDAS as we expect these outcomes to take longer than the duration of these trials. We did observe an effect for the secondary outcome of decrease in lesion size.

The clinical implications of these results is uncertain due to moderate to high risk of bias. The two studies that were masked and had moderate risk of bias showed no effect, while the two studies with high risk of bias and lack of masking supported the caries arrest outcome. It is possible to mask assessors (and all providers, patients, and statisticians) in trials of CR. Researchers can and should conduct masked CR trials that are longer, larger, and in populations with higher caries activity. Nonetheless, non-masked studies have led to important progress in oral health care, such as SDF affecting the patient-centered outcome of avoiding general anesthesia.^45–47^

Two studies contributing to decreased lesion size results had masked assessment.^35,36^ Absolute size from one trial^36^ was converted to relative size to match the scale of the other and thereby enable combination through meta-analysis. Focus on relative change brings limitations,^48^ but the measurement technique in the other study^35^ did not enable conversion to the absolute size needed to assess the contrast effect.

These limitations are similar to those for the studies included in the systematic review and network meta-analysis underlying the ADA Center for Evidence-based Dentistry clinical practice guidelines for non-restorative caries treatment,^18,31^ which has been the standard US guidance for non-invasive dentistry since 2018.

As there are tragically few therapeutic agents for dental caries, it is exciting that there are ten clinical trials demonstrating an anti-caries effect by CR.^33–36,39,40,44,49–51^ Clinical trials supporting other brush-on treatments for caries include those of traditional fluorides, sealants (including resin-infiltration), and SDF (including silver nitrate and fluoride varnish).^18,52^ This is perilously insufficient, considering that caries is the most common disease in humans. The only alternative treatments are technique-sensitive, expensive, and invasive operative procedures that require maintenance and repeated replacement over time.

### Implications for research

This meta-analysis suggests that longer and larger trials may show effects across cavitation and decrease in merged ICDAS. We expect clinical trials of dental caries to last 18 to 24 months.^53–56^ However, caries arrest trials can show an effect after just two weeks.^57^ The trials here ranged 6 to 12 months. Longer trials are needed to discern long-term outcomes, assess prevention of progression, and assess whether re-application is beneficial or necessary to maintain or build effect over time, as it is with SDF.^58^

The combination of CR and simultaneous fluoride might be synergistic. Two trials combining fluoride varnish with CR trended toward a higher effect than those with no fluoride. All control groups in trials that reported using ICDAS criteria received fluoride varnish. The group treated with CR only performed worse than fluoride varnish, while groups treated with both CR and fluoride varnish trended better than fluoride varnish only. Therefore, we may expect enamel regeneration incorporating fluoride to be more successful than not. Future research to determine whether CR acts synergistically with fluoride varnish may do so by comparing CR to CRFP.

The studies on occlusal lesions showed a strong effect on caries arrest. The studies showing an effect on lesion size were on facial and/or buccal lesions. We recommend conducting studies to determine the respective effects on the converse surfaces, and on approximal surfaces.

### Implications for practice

The positive effect on promoting caries arrest and decreased lesion size suggests judicious clinical use of CR for initial caries lesions. This finding is a clinically meaningful addition beyond the effect of preventive behavior change and other interventions such as fluoride toothpaste and fluoride varnish. There may be an additive or synergistic effect when combined with fluoride varnish, so it is a potentially valuable addition compared to fluoride varnish alone.

International guidance for caries management has been building for some time. The International Caries Classification and Management System (ICCMS) Guide^59^ brought together 75 authors to distill best evidence into recommendations supported by a Policy Statement from the FDI World Dental Federation. The *CariesCare International* practice guide^60^ repackages the ICCMS Guide into a practice-friendly format that engages patients as long-term health partners and uses a ‘4D cycle’: Determine risk, Detect disease, Decide on a personalized care plan, and Do the provision of personalized tooth-preserving care, including control of initial lesions. Caries management by CR fits well within this overall care framework.

Expanding the number of available non-invasive treatment options for initial lesions also has the promise of improving outcomes. For example, a study of a school program of dental hygienists applying SDF, sealants, fluoride varnish, 10% povidone iodine, and temporary restorations without excavation found a 69% decrease in treatment of caries under general anesthesia.^46^ Considering that over 90% of dentists report routinely restoring initial caries lesions,^61,62^ despite international guidelines concluding that this does more harm than good,^63^ having another non-invasive alternative would benefit patients. There is also a range of economic benefits from re-orientating oral health services towards non-invasive care.^64^ If funded at a lower rate than restorations and performed by non-dentist dental team members, it may be straightforward to create reimbursement strategies that are beneficial to payers, dental teams, and patients.

The product currently available in the US (CRFP) is identical to the international product except it contains 500 ppm sodium fluoride. Therefore, the generalizability of these results within the US is unknown. While NMR analysis shows that this does not have a substantial effect on fibril assembly (unpublished data from the manufacturer), it may impact clinical effect, which could be negative or positive.

## Conclusions

This systematic review and meta-analysis provide evidence that CR is effective in arresting initial (non-cavitated) caries lesions across four studies and reducing lesion size across two studies. Further research is needed to clarify the effects on preventing cavitation and merged ICDAS. All six included trials have moderate to high risk of bias. Longer trials with low risk of bias are recommended. Curodont™ Repair is an intriguing addition to the limited pharmacopeia for the most common disease in humans, dental caries.

## Data Availability

All data produced in the present study are available upon reasonable request to the authors.

## Figure and Table Legends

**Supplemental Table 1.** Details of the Risk of Bias analysis

**Supplemental Figure 1.** Subgroup analysis on both 1. the effect of fluoride varnish in both CR and control groups at the baseline intervention and 2. patient-randomization (P_11_-4 + fluoride) versus split-mouth (P_11_-4 only) on caries arrest. These two factors result in the same delineation between studies.

**Supplemental Figure 2.** Subgroup analysis on the effect of study duration (≤6 versus >6 months) on caries arrest.

**Supplemental Figure 3.** ICDAS outcomes for Curodont Repair group, from Kobeissi et al., 2020. In the published paper the respective result for the fluoride varnish group was errantly duplicated for both.

**Supplemental Figure 4.** Meta-analyses of decrease in merged-ICDAS score in randomized-controlled trials of CR by risk difference.

