## Supplementary Table 1 for "Systematic Review and Meta-Analysis on the Effect of Self-Assembling Peptide P_11_-4 on Initial Caries Lesions"

| Risk of Bias (ROB) | Alkilzy | Gözetici | Doberdoli | Sedlakova Kondelova | Welk | Kobeissi |
|---|---|---|---|---|---|---|
| Study design and details | Parallel arm RCT. Duration = 6 months. 70 patients, 70 lesions. Average age = 10.0 years. Groups: 1. Curodont, 2. no treatment. Both groups received F varnish at D0 & D90. | 4 way split mouth RCT. Duration = 6 months. 21 patients, 84 lesions. Average age = 15.4 years. Groups: 1. Curodont, 2. Icon, 3. F varnish, 4. no treatment. | Parallel arm RCT. Duration = 12 months. 90 patients, 90 lesions. Average age = 11.8 years Groups: 1. Curodont, 2. Curodont + pre-assembled Curodont at home, 3. no treatment. Groups 1 & 3 had F varnish at D0 & D180. | Split mouth RCT. Duration = 9 months. 44 patients, 88 lesions. Average age = 27.1 years. Groups: 1. Curodont, 2. Placebo. Both groups received F varnish at D90. | Split mouth RCT. Duration = 6 months. 23 patients, 46 lesions. Average age = 15.4 years. | Split mouth RCT. Duration = 6 months. 9 patients, 40 lesions. Average age = 11.1 years. Groups: 1. Curodont, 2. F varnish |
| ROB arising from the Randomization process: | Some - 3rd party comp-generated random allocation sequence. Laser fluorescence and VAS scores showed test group lesions were larger at baseline; but grouped ICDAS baseline scores were similar. | Some - Quasi-randomization: Randomization by 3rd party, but if there was more than one tooth in the quadrant, investigators "selected one" The control group had the lowest diagnodent scores. | Low - Good randomization by 3rd party. No baseline differences identified. | Low - Good randomization by 3rd party. No baseline differences identified. | Some - Randomization = flipping a coin. Baseline difference of 8.8 $mm^2$ (test) vs 6.8 $mm^2$ (control) in morphometric measurement. Baseline impedance values were very similar. | Low - Simple randomization by flipping a coin. Grouped ICDAS baseline scores were similar. |
| Adherence: ROB due to assignment to intervention, intention to treat and "blinding": | Some - The study was unblinded. No placebo experience. | Some - The treatment investigators were unblinded. The patients had 4 treatments – so likely blinded. | Some - The treatment investigators were unblinded. No placebo experience; single treatment and outcome investigator. | Good - Quadruple blinded; placebo experience. | Some - No placebo experience. Unblinded treatment investigator. | Some - The study was unblinded. No placebo experience. |
| Missing Outcome Data: | Low - 8 patients (11%) were lost. 62/70 patients at endpoint. | Low - 1 patient (5%) was lost. 20/21 patients at endpoint. | Low - 8 patients (13%) were lost. 52/60 patients at endpoint. | Low - 4 patients (9%) were lost. 40/44 patients at endpoint. | High - 2 patients (9%) were lost. 21/23 patients were seen at endpoint. But only 14/23 patients (61%) could be assessed for morphometry at endpoint. | Low - 9/9 patients at endpoint. |
| ROB due to Measurement of the outcome: | High - No report that the outcome assessor(s) were blinded. (Number of assessors unknown). | Low - The single assessor was blinded and different from the allocation investigator. | High - One single investigator for application and evaluation. | Low - Blinded outcome assessor. | Low - Blinded outcome assessor & statistician. | High - unblinded and subjective assessment. Unpredictable direction in outcome measurement. |
| ROB in selection of the reported result: | Some - Trial registration after study completion. | Some - No trial registration. | Low - Trial registration after study completion. Yet very fair and logical reporting. | High - Trial registration after study completion. Illogical timelines and group comparisons in report. | High - No trial registration nor statistical analysis preplan reported. | Some - No trial registration. |
| Overall ROB | High - Due to unblinded outcome assessors and company sponsorship. Note: self-report, "No role in study design, data collection, data analysis, data interpretation, or writing of the report." | Some - Company supplied Curodont. Risk of bias direction is unpredictable, results section promote resin infiltration and fl varnish, not Curodont, despite positive outcomes. | High - Due to unblinded, single investigator. Company sponsored and co-authored. | Some - Good blinding, but creative reporting. Company supplied material and co-author. | High - Due to creative outcome reporting and missing outcome data. | High - Due to unblinded assesssment, though direction unpredictable. Reported sources of support and conflicts of interest are "none." |
