## Supplementary figures and images for "Systematic Review and Meta-Analysis on the Effect of Self-Assembling Peptide P_11_-4 on Initial Caries Lesions"

### Figure S1

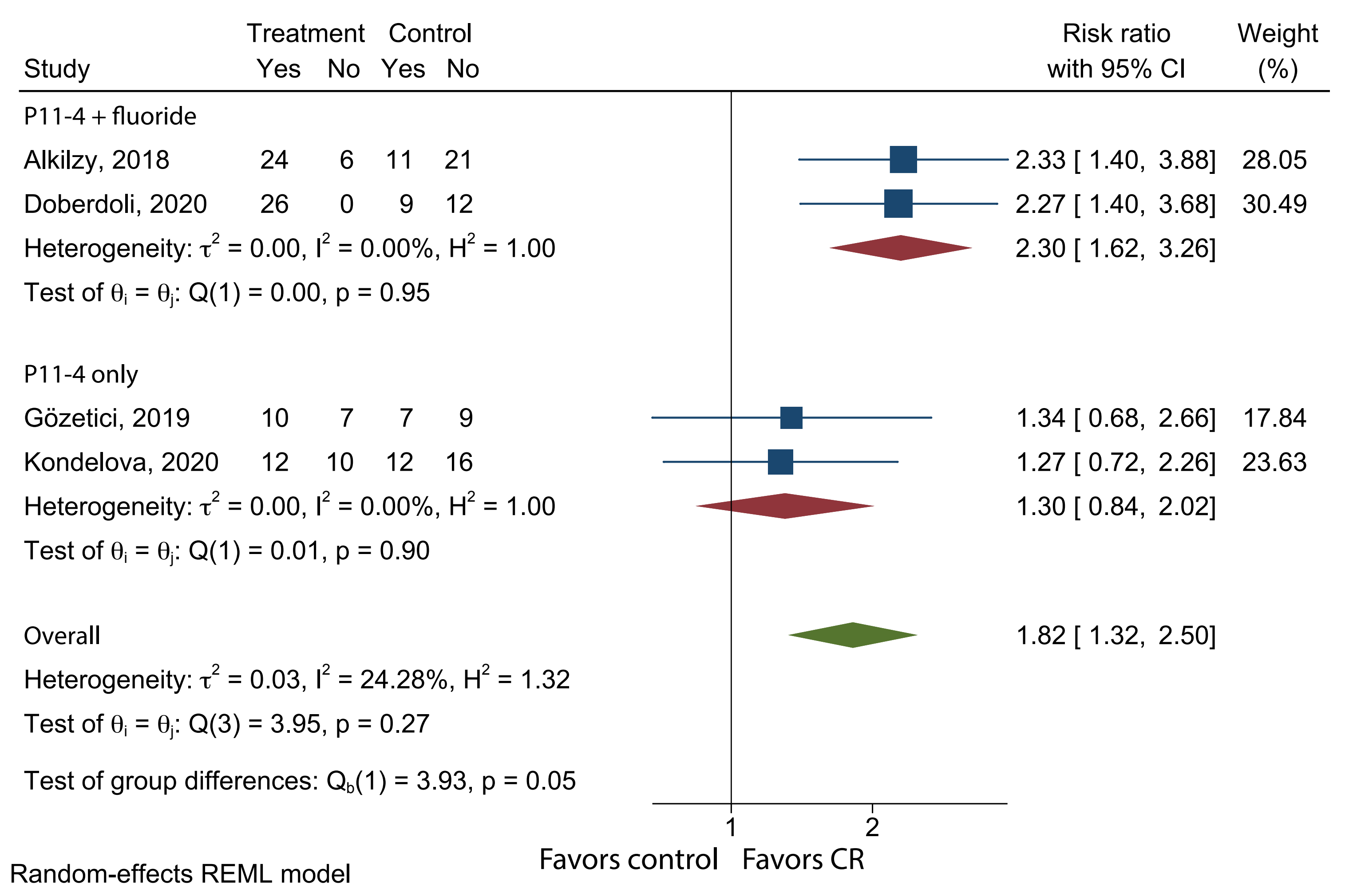

### Figure S2

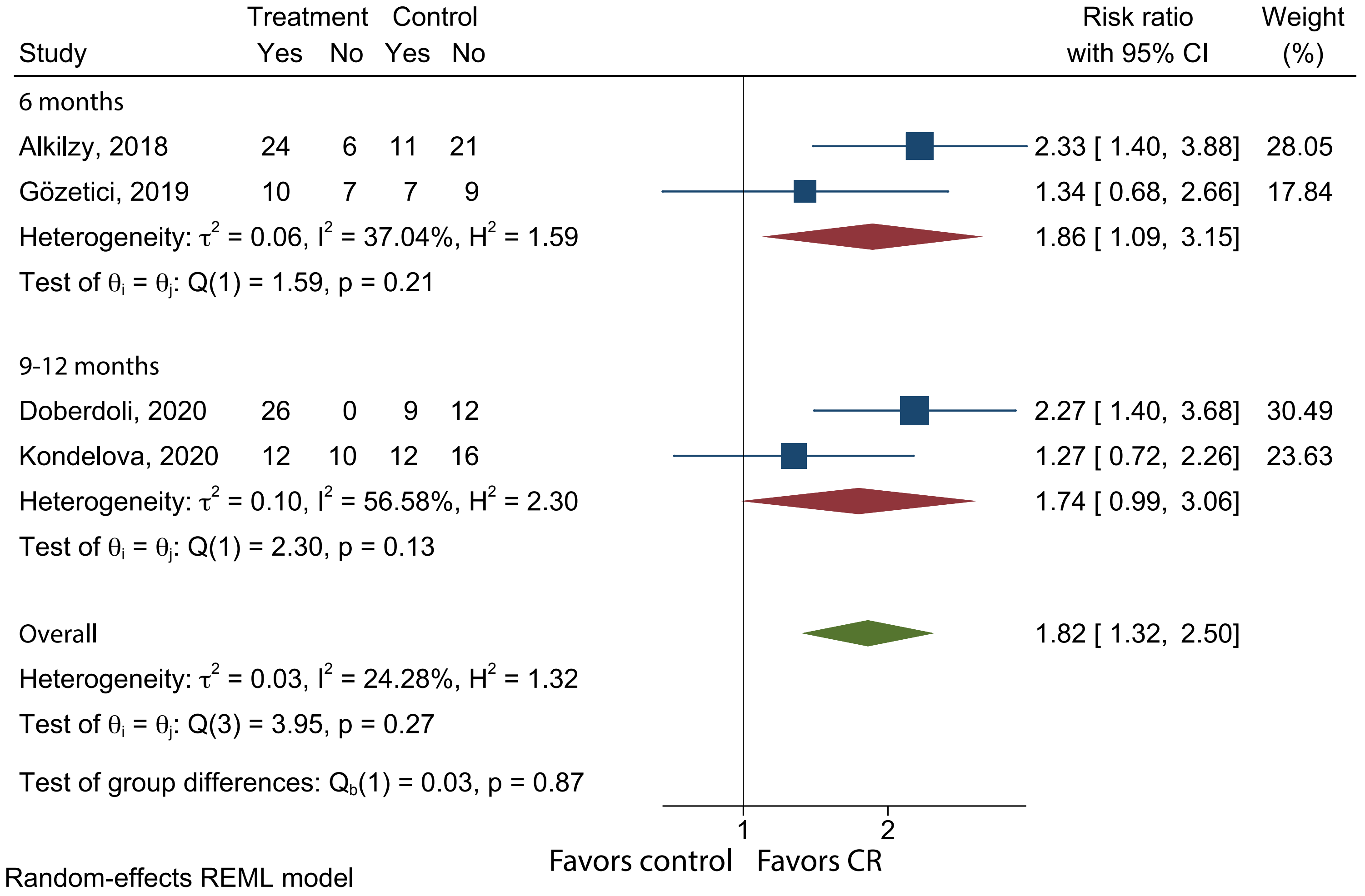

### Figure S3

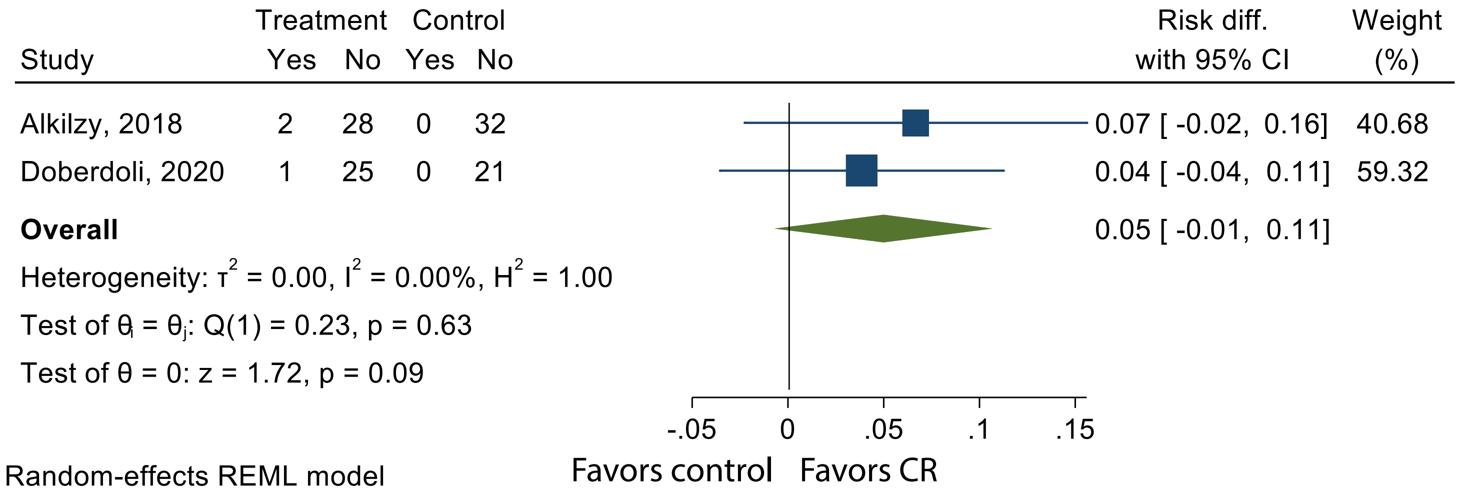

### Figure S4

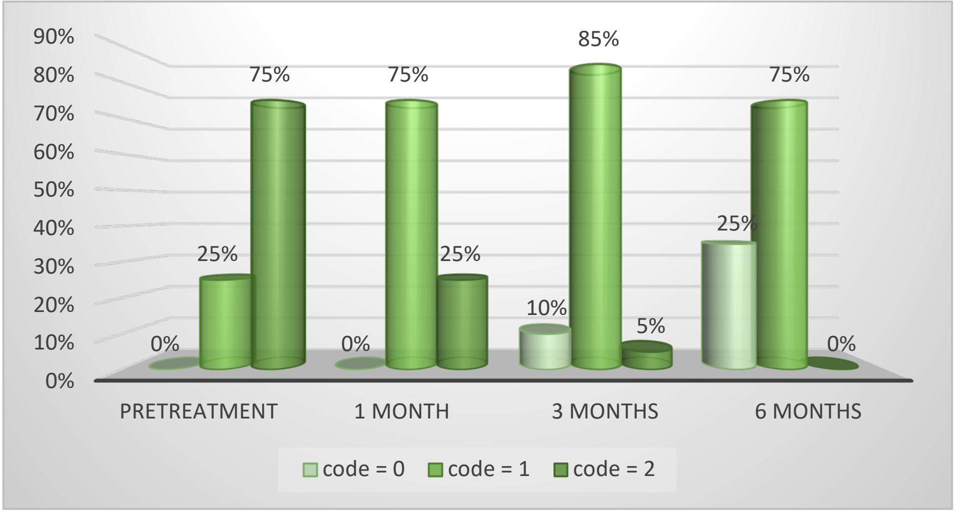
